# A calibrated temporal reference map of disease progression

**DOI:** 10.64898/2026.06.24.26356443

**Authors:** Jiazi Tian, Alaleh Azhir, Jonas Hügel, Chirag J. Patel, Hossein Estiri

**Author notes:** Correspondence: [ ].

## Abstract

**Background:** Understanding the evolution of human illness requires capturing the temporal directionality of disease progression, yet existing biomedical reference maps largely describe cross-sectional states or static comorbidity. We introduce a directed, probability-ranked map (i.e., a knowledge-base) of clinical progression derived from population-scale longitudinal electronic health records.

**Methods:** The knowledge-base was constructed from de-identified EHRs of 295,678 individuals across the Mass General Brigham system, yielding 435,240 phenotype-pair-duration associations via temporal Spearman correlation. To distinguish biological progression from administrative artefact at scale, we distilled a locally deployed MedGemma labeling function into two complementary classifiers: a RF capturing local episodic signal and a GNN aggregating global network topology via message passing. Their outputs were combined as an unweighted late-fusion average. Classifier confidence was systematically evaluated against pairwise genome-wide genetic correlation estimates from the UK Biobank as an independent biological reference standard.

**Results:** Both classifiers achieved comparable distillation fidelity on the 200-row development set (RF AUROC 0.772; GNN AUROC 0.769). Genetic support was concentrated in the highest confidence deciles, with both models achieving highly significant top-decile enrichment for validated genetic pleiotropy (RF: 1.36-fold, *p* < 0.001; GNN: 1.32-fold, *p* < 0.001), demonstrating that classifier confidence aligned with independent genomic support. The framework additionally identified two complementary classes of progression: acquired mechanical cascades with high classifier confidence but null genomic overlap (exemplified by musculoskeletal pain progressing to cardiac dysrhythmias beyond 90 days, 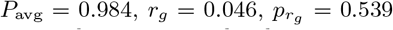), and topological bridges structurally enforced by network architecture despite sparse local co-occurrence (exemplified by acute myocardial infarction to epilepsy within 0–14 days, *P*_GNN_ = 0.930 versus *P*_RF_ = 0.332).

**Conclusions:** By transitioning from static comorbidity networks to a confidence-ranked landscape of temporal trajectories, the map provides a biologically calibrated coordinate system for prioritising mechanistic, translational, and clinical investigation of disease progression.

## 1 Introduction

Foundational biomedical reference maps have transformed modern biology by organizing genes, tissues, and cells into shared coordinate systems that enable systematic discovery. Disease relationships have similarly been mapped through molecular interaction networks, comorbidity analyses, and population-scale association studies, revealing clusters of shared risk and overlapping genetic architecture [1, 2]. Yet most existing disease maps are inherently cross-sectional: they characterize similarity, co-occurrence, or shared susceptibility without explicitly representing how illness unfolds over time.

Understanding disease evolution requires not only identifying which conditions are related, but also determining the temporal directionality through which one clinical state transitions into another. A reference framework for disease progression, analogous to molecular atlases but temporally resolved, remains largely absent. Evolutionary biology resolved a related problem with phylogenomics, the reconstruction of evolutionary relationships from genome-scale data [3]. Its organizing unit is the clade, a group that contains one ancestor and all of its descendants [4]. Clades are monophyletic and nested, so each branch splits into finer branches, and the structure records the order in which lineages diverged. The discipline reads biology through directed, branching descent rather than through static similarity. We borrow that structure and call the missing clinical framework the *phylophenome*: a directed, time-resolved map of how clinical phenotypes follow one another across a population. The genome catalogues heritable variation and the proteome catalogues expressed proteins; the phylophenome catalogues the order in which disease states succeed one another. A clinical clade is one resolved trajectory through that map, the path from a baseline state to the complications that branch from it. Phenotypes do not inherit from one another in the Darwinian sense, yet the directed, branching topology of the clade describes their temporal succession well.

Longitudinal electronic health records (EHRs) provide a unique opportunity to address this challenge by capturing sequential patterns of diagnosis across large populations and extended periods of observation [5, 6]. Prior studies have leveraged EHR data to model trajectories of multimorbidity and identify temporal disease associations [5, 7, 8]. However, distinguishing biologically meaningful progression from observational artefact remains a major unresolved challenge. Healthcare-derived temporal associations are shaped not only by disease biology, but also by coding practices, referral pathways, institutional workflows, delayed diagnoses, and iatrogenic processes [9]. Consequently, statistically significant temporal correlations may reflect administrative structure rather than genuine mechanisms of disease progression. Although expert adjudication through chart review can identify plausible transitions, manual review cannot scale to the hundreds of thousands of candidate trajectories generated in contemporary healthcare systems.

Recent advances in biomedical large language models offer a potential mechanism for scaling expert-like clinical reasoning across observational data [10, 11]. Rather than treating temporal association as sufficient evidence of progression, language models may provide a means to algorithmically triage candidate trajectories according to mechanistic plausibility. At the same time, disease progression is inherently multi-scale: some transitions are supported by dense local temporal evidence, whereas others emerge only through broader network context. Capturing these complementary structures requires integrating both local episodic signal and higher-order relational topology.

A second challenge lies in establishing trust in progression maps derived from a single healthcare system. Observational models risk encoding local institutional behaviour rather than generalisable biology [9]. Orthogonal biological validation is therefore essential. Genome-wide genetic correlation quantifies the extent to which two traits share heritable architecture and provides an independent measure of biological relatedness derived from genome-wide association studies [12]. Because these estimates are derived from independent population-scale genetic studies rather than local healthcare workflows, concordance between inferred clinical trajectories and shared genetic architecture may provide an orthogonal measure of biological plausibility [12, 13].

Here, we introduce a biologically calibrated map (i.e., a knowledge-base) of disease evolution, derived from longitudinal electronic health records across 295,678 individuals within a large integrated healthcare system. This map is our first instantiation of the phylophenome, built from a single integrated health system to preserve temporal continuity. The knowledge-base represents phenotypes as directed temporal transitions ranked by probabilistic confidence, transforming large-scale observational records into a temporal coordinate system for clinical progression. To enable scalable adjudication, we distil expert-style biomedical reasoning from a locally deployed language model into complementary predictive frameworks that integrate local temporal structure and global network topology. We then evaluate the resulting progression landscape against independent genome-wide genetic correlation estimates from the UK Biobank as an orthogonal measure of biological support. Beyond recovering trajectories enriched for shared heritable architecture, we show that systematic discordance between temporal progression and genomic overlap reveals acquired, mechanically mediated pathways that conventional genome-wide approaches fail to capture.

## 2 Results

### 2.1 Study Population and Knowledge-base Construction

The knowledge-base was derived from de-identified EHRs across the Mass General Brigham (MGB) system spanning 2015–2023. The cohort comprised 295,678 patients in three groups: COVID-19 cases (*n*=85,364), post-pandemic controls (*n*=170,497), and pre-pandemic controls (*n*=39,817). Demographic and clinical characteristics were balanced across groups by design (Table 1). EHR records were mapped to the Clinical Classification Software Revised (CCSR)[14] before we distilled all phenotype pairs from the EHR records using transitive sequential patterns mining (tSPM+) [15] analogue to [16]. Based on the binned duration, i. e. the temporal distance between the occurrence of the single phenotypes, of phenotype pairs (0-14 days, 15 days to 30 days, 31 to 90 days, 90+ days) we computed temporal Spearman correlations for all CCSR phenotype pairs across the four duration bins, yielding a reference corpus of 435,240 temporal associations.

**Table 1:** Study population characteristics. The same cohorts were used in a previous study [16] to develop an algorithm to identify patient with specific post-COVID symptoms.

| Characteristic | COVID-19 Cases<br>($n=85,364$) | Post-Pandemic Controls<br>($n=170,497$) | Pre-Pandemic Controls<br>($n=39,817$) |
| --- | --- | --- | --- |
| Age, mean (SD) | 53.6 (17.2) | 53.7 (17.3) | 54.7 (17.5) |
| Female, % | 62.6 | 62.5 | 63.2 |
| Charlson index, mean | 2.2 | 2.2 | 2.0 |
| White, $n$ (%) | 60,935 (71.4) | 122,646 (71.9) | 29,687 (74.6) |
| Black, $n$ (%) | 8,409 (9.9) | 16,688 (9.8) | 3,273 (8.2) |
| Hispanic ethnicity, $n$ (%) | 6,110 (7.2) | 8,840 (5.2) | 1,950 (4.9) |
| Date range | 2017–2023 | 2017–2023 | 2015–2020 |

### 2.2 Classification Performance on the Development Set

Both the Random Forest (RF) and Graph Neural Network (GNN) were trained on 10,000 MedGemma-labeled sequences and evaluated against the 200-row Labeling Function Development Set. Because this set shaped the prompt that generated the training labels, performance on it quantifies algorithmic distillation within the development loop rather than out-of-sample generalisation.

Table 2 reports the metrics. The MedGemma labeling function itself achieved Cohen’s *κ* = 0.508 against the human consensus on this set (LLM baseline row, Table 2), establishing the silver-standard ceiling toward which the RF and GNN were distilled. The RF achieved a training AUROC of 0.802 (5-fold cross-validated AUROC 0.806 ± 0.008) and a development-set AUROC of 0.772. The GNN achieved a development-set AUROC of 0.769. The near-identical development-set AUROCs reflect convergent distillation fidelity from architecturally distinct models; their scientific value lies in complementarity rather than relative performance. Given the class imbalance (133 NON-NOISE to 67 NOISE instances), secondary metrics calibrated for skewed prevalence are additionally reported, including the Matthews Correlation Coefficient and balanced accuracy. Both classifiers learned the decision boundaries defined by the LLM without collapsing onto the majority class.

**Table 2:** Distillation fidelity on the human-curated development set (*n* = 200).

| Model | Sens. | Spec. | AUROC | PPV | NPV | F1 | MCC | Acc. |
| --- | --- | --- | --- | --- | --- | --- | --- | --- |
| LLM baseline (MedGemma) | 0.786 | 0.739 | — | 0.851 | 0.646 | 0.818 | 0.511 | 0.770 |
| Random Forest | 0.748 | 0.710 | 0.772 | 0.831 | 0.598 | 0.787 | 0.443 | 0.735 |
| Graph Neural Network | 0.718 | 0.696 | 0.769 | 0.817 | 0.565 | 0.764 | 0.397 | 0.710 |

Sens., sensitivity; Spec., specificity; AUROC, area under the receiver operating characteristic curve; PPV, positive predictive value; NPV, negative predictive value; F1, harmonic mean of precision and recall; MCC, Matthews correlation coefficient; Acc., accuracy. AUROC is undefined for the MedGemma labeling function because it produces deterministic binary labels rather than calibrated probabilities.

### 2.3 Biological Enrichment and Pleiotropic Recovery

We tested whether classifier confidence prioritises sequence pairs with shared genetic architecture, providing an external biological validity check independent of the development set. Predictions were stratified into ten confidence deciles and the prevalence of validated genetic pleiotropy 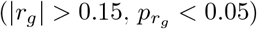 was computed within each decile (*N* = 1,061 independent phenotype pairs; permutation test, 1,000 iterations).

Figure 1 shows the decile distributions for both classifiers. Against a baseline prevalence of 63.0%, the RF produced 85.8% prevalence in the top confidence decile (1.36-fold enrichment, *p* < 0.001). The GNN produced 83.0% in the top decile (1.32-fold, *p* < 0.001). Both classifiers showed suppression below baseline in the lowest decile (RF: 42.1%; GNN: 39.3%), indicating bidirectional discrimination at the extremes of confidence. Genetic support was elevated at the highest confidence strata and depleted at the lowest, demonstrating that classifier confidence serves as a quantitative proxy for genomic concordance.

**Figure 1:**
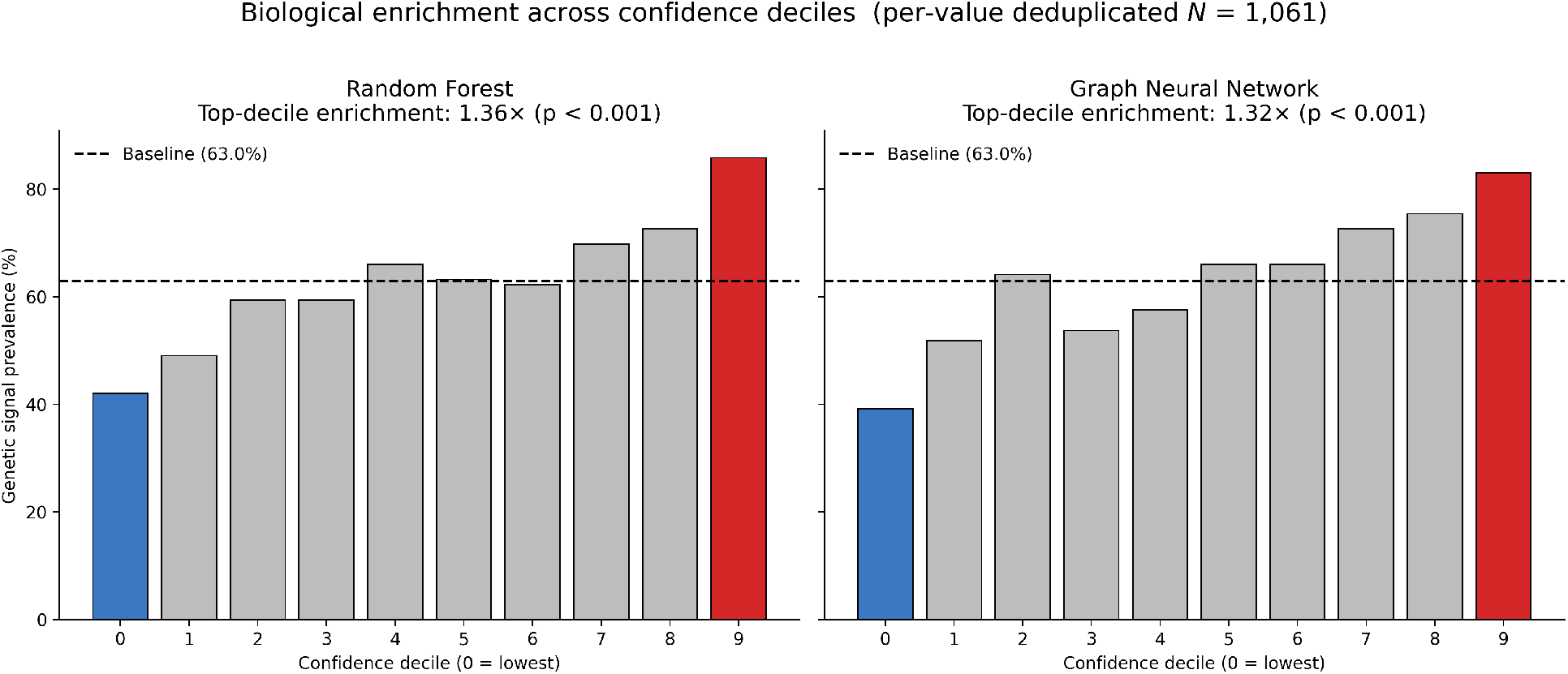
Biological enrichment across confidence deciles. Prevalence of validated genetic pleiotropy 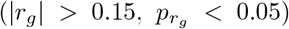 across ten deciles of classifier confidence, on a per-value deduplicated set of independent phenotype pairs (*N* = 1,061). The RF top decile reaches 85.8% prevalence (1.36-fold over the 63.0% baseline, *p* < 0.001). The GNN reaches 83.0% in the top decile (1.32-fold, *p* < 0.001). Both classifiers show suppression below baseline in the lowest confidence decile (RF: 42.1%; GNN: 39.3%). The elevation of genetic support in the highest strata and its depletion in the lowest demonstrate that the averaged classifier confidence serves as a biologically calibrated ranking of disease transitions.

### 2.4 Case Study: Unsupervised Mechanistic Discovery

#### Candidate Non-Pleiotropic Temporal Progression

Genome-wide genetic correlation does not capture the full spectrum of longitudinal clinical trajectories observed in routine care. To identify high-confidence progressions that may operate independently of shared polygenic susceptibility, we filtered the adjudicated sequence matrix for extreme late-fusion confidence (*P*_avg_ > 0.95) alongside strictly confirmed null genomic correlation 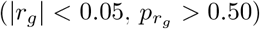. This prioritisation strategy isolates candidate acquired, post-acute, or mechanically mediated pathways that may not be detectable through shared heritable architecture alone.

Using this framework, the Temporal Knowledge Base (TKB) identified a progression from musculoskeletal pain (excluding low back pain) to cardiac dysrhythmias at a temporal lag exceeding 90 days. The latefusion average assigned this trajectory a probability of 0.984 (*P*_RF_ = 0.980, *P*_GNN_ = 0.989), with strong support from the underlying longitudinal EHR signal (*ρ* = 0.510, *p* = 3.8 × 10^−79^). UK Biobank genetic correlation estimates for this phenotype pair were statistically null 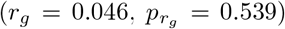, suggesting limited evidence for shared polygenic liability. Although the present analysis cannot establish causality, this transition is consistent with clinically plausible autonomic mechanisms in which persistent nociceptive burden may contribute to sustained sympathetic activation and increased arrhythmic vulnerability. The TKB therefore prioritises a candidate temporal progression that would be poorly captured by genetic overlap alone (Figure 2).

**Figure 2:**
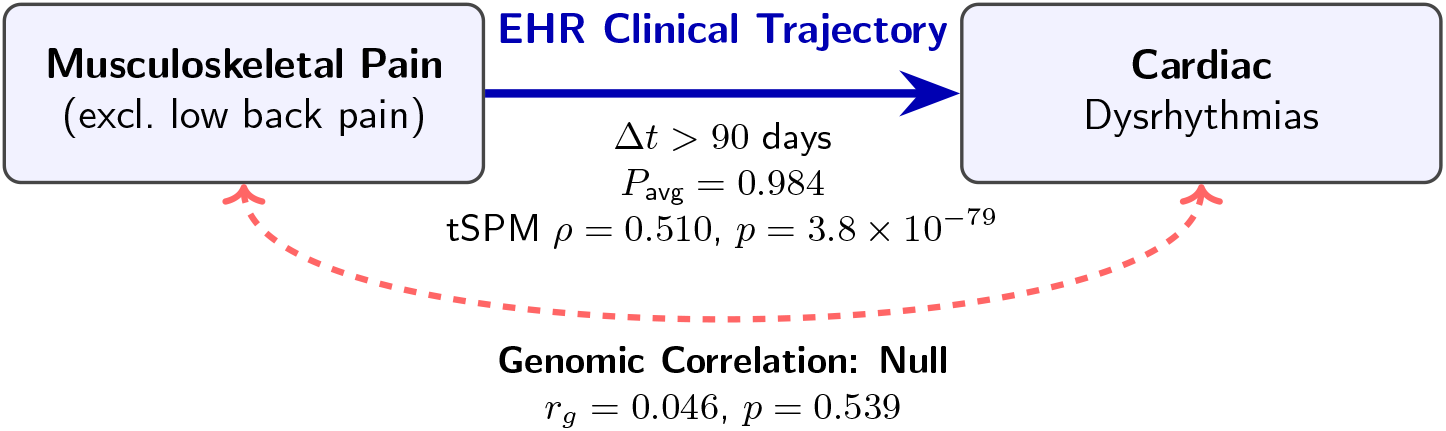
Candidate non-pleiotropic temporal progression. Schematic representation of the temporal progression from musculoskeletal pain to cardiac dysrhythmias at Δ*t* > 90 days. The high late-fusion confidence (*P*_avg_ = 0.984, *P*_RF_ = 0.980, *P*_GNN_ = 0.989) and strong EHR-derived temporal signal (*ρ* = 0.510, *p* = 3.8× 10^−79^) prioritise this trajectory despite null genomic overlap (*r*_*g*_ = 0.046, *p* = 0.539). The absence of detectable genetic correlation suggests a progression that may reflect acquired or mechanically mediated processes rather than shared polygenic susceptibility.

#### Topology-Supported Temporal Transition

Systematic divergence between the two constituent models identified candidate transitions that were weakly represented by local co-occurrence but supported by broader network topology. We therefore searched the directed network for sequences characterised by low RF confidence but high GNN confidence, reflecting potential structural reinforcement through neighbourhood context.

The progression from acute myocardial infarction to epilepsy and convulsions within a 0–14 day temporal window yielded a low RF probability of 0.332, consistent with sparse direct temporal co-occurrence in the local training data. In contrast, the GNN assigned a high probability of 0.930, indicating strong topological support derived from the surrounding phenotype network. The underlying EHR association was modest but statistically significant (*ρ* = 0.098, *p* = 7.3 × 10^−4^), while UK Biobank genetic correlation demonstrated substantial shared heritable architecture 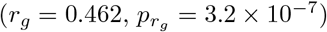.

Although the present framework cannot establish mechanistic causality, this acute transition is consistent with known cardiovascular–neurological associations reported in prior clinical studies.[17] This example illustrates the complementary role of the two classifiers: the RF preferentially captures locally dense temporal signal, whereas the GNN recovers transitions supported by broader network structure that may be difficult to identify through isolated co-occurrence statistics alone (Figure 3).

**Figure 3:**
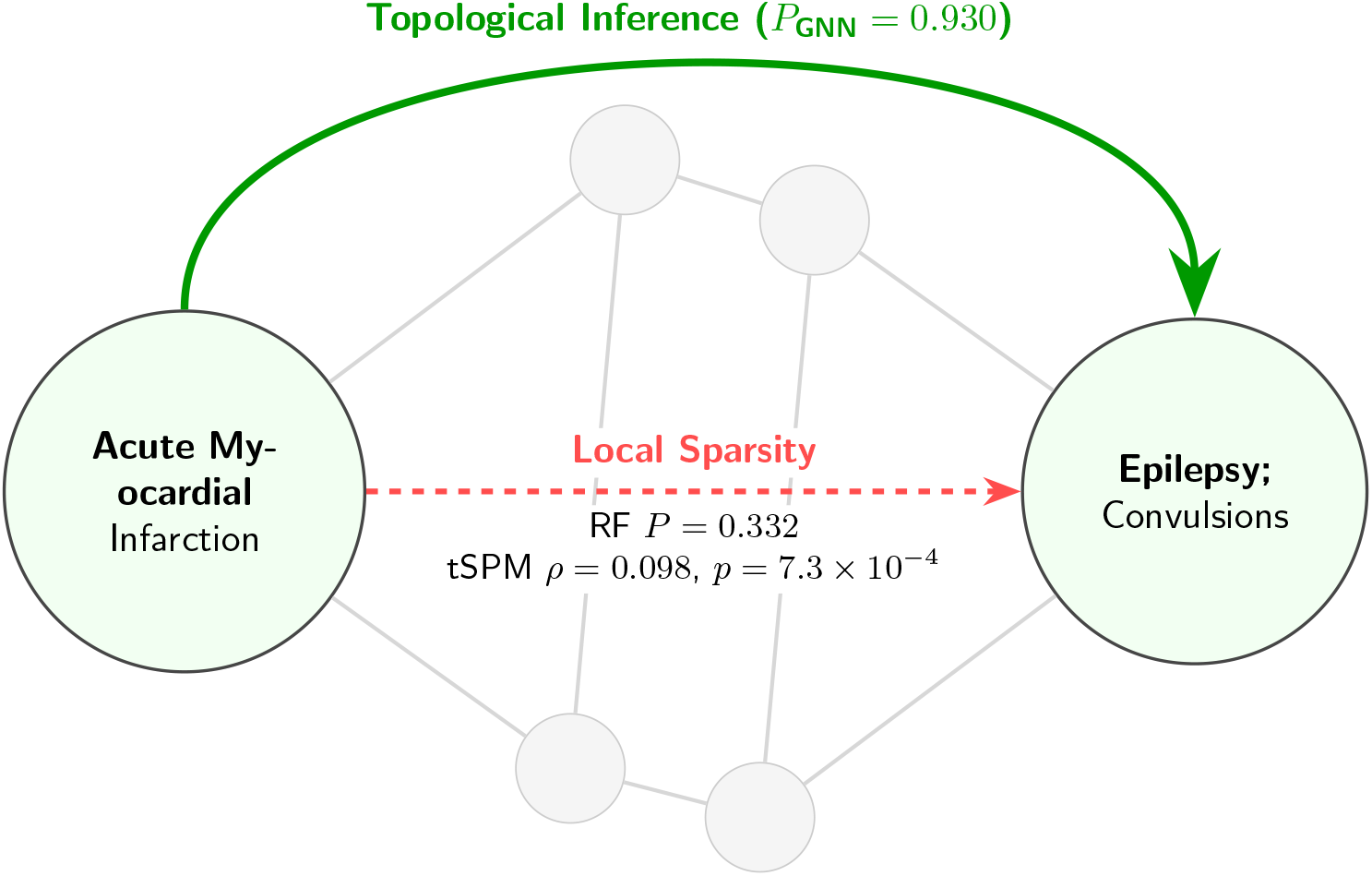
Topology-supported temporal transition. The RF assigns low confidence (*P*_RF_ = 0.332) to the acute myocardial infarction → epilepsy transition within the 0–14 day window because of sparse direct temporal co-occurrence. The GNN assigns high confidence (*P*_GNN_ = 0.930) by incorporating surrounding network topology through message passing. Although the EHR-derived temporal association is modest (*ρ* = 0.098, *p* = 7.3× 10^−4^), strong genetic overlap (*r*_*g*_ = 0.462, *p* = 3.2 ×10^−7^) supports a broader shared biological relationship. This example illustrates how topological context may prioritise clinically plausible temporal transitions that isolated pairwise statistics under-emphasise.

## 3 Discussion

Existing disease maps largely describe similarity, co-occurrence, or shared molecular architecture [1]. While most biomedical reference maps describe state, this TKB describes transition. The knowledge-base is not merely a disease network; it is a temporal coordinate system for disease evolution. By applying multi-modal adjudication to half a million clinical sequences, the TKB reconstructs large-scale patterns of disease progression observed in longitudinal clinical practice. This temporal organization reveals progression pathways, transition bottlenecks, and bridge conditions that remain hidden in static, cross-sectional networks, providing a framework for studying how diseases evolve over time.

A defining feature of this foundational knowledge-base is its biological validation against orthogonal genomic evidence. Both classifiers successfully recovered progressions with shared heritable architecture, achieving highly significant top-decile enrichment for validated genetic pleiotropy (85.8% and 83.0% prevalence, respectively; *p* < 0.001). However, the most striking observation was not simply enrichment among the highest-confidence trajectories, but the alignment between classifier confidence and independent biological support across the range of confidence. Genetic support was concentrated among the highest-confidence trajectories and systematically depleted in the lowest-confidence predictions. This graded pattern suggests that the ranking structure of the network itself captures biologically meaningful information. The late-fusion average confidence score serves not merely as a computational metric, but as a biologically calibrated ranking of disease transitions.

Unlike traditional disease networks that represent relationships as present or absent, the TKB provides a confidence-ranked landscape of disease progression. The observed alignment between confidence and genetic support suggests that transition probabilities capture more than statistical association. Instead, they provide a quantitative estimate of biological plausibility that can be used to prioritize downstream mechanistic, translational, and clinical investigation. In this sense, the knowledge base functions not only as a map of disease progression but also as a framework for allocating scientific attention.

A central contribution of the TKB is the identification of topological bridges, transitions that are weakly represented by direct temporal co-occurrence yet become apparent when viewed within the broader structure of disease progression. For example, the progression from acute myocardial infarction to epilepsy within a 0–14 day window features sparse local co-occurrence but high structural connectivity within the broader network. While strong genomic overlap supports a shared long-term cardiovascular-epilepsy axis, the knowledge-base isolates the acute mechanistic bridge. Plausible acute transitions, such as post-MI cardioembolic micro-infarcts lowering the seizure threshold, are structurally enforced by the surrounding network topology independently of the polygenic background.

Crucially, perfect concordance between clinical sequence probability and genetic correlation is neither expected nor biologically plausible. While genome-wide association studies map shared polygenic susceptibility, the electronic health record captures acquired, environmental, and mechanical cascades. By filtering for extreme late-fusion confidence alongside strictly null genetic correlation, the knowledge-base systematically isolates pathways operating via systemic damage rather than shared heritability. The discovery of the musculoskeletal pain to cardiac dysrhythmias trajectory demonstrates this utility. This cascade is consistent with acquired autonomic mechanisms, such as chronic nociceptive pain driving sustained sympathetic activation, that leave no pleiotropic trace [18]. The discordance between the knowledge-base and genomic overlap is therefore a feature that systematically maps non-genetic biological transitions.

A standing critique of observational clinical models is their reliance on single-system healthcare data. This limitation reflects a deliberate trade-off between scale and temporal continuity. Integrated healthcare systems provide continuous longitudinal observation, whereas federated networks and claims databases offer broader geographic representation. Because transition discovery depends on the precise sequencing of clinical events, we prioritized temporal continuity. The inherent heterogeneity in institutional coding practices and fragmented follow-up within massive federated networks may obscure the strict temporal fidelity required to capture acute clinical transitions. Furthermore, validating temporal trajectories from a regional cohort against the biological architecture of an orthogonal, international genome-wide dataset supports the interpretation that many high-confidence trajectories reflect underlying biological relationships rather than local administrative artifacts.

What computational inference across observational data cannot definitively prove is strict causality. The TKB operates as an engine to narrow the search space for mechanisms, not as a substitute for targeted molecular biology or prospective clinical validation. A primary limitation of this study is the necessary coupling of our human-adjudicated development set to the prompt-generation process, preventing its use as a strictly held-out test set for final expert concordance. Future extensions incorporating additional healthcare systems, genomic resources, and prospective validation cohorts may enable increasingly comprehensive maps of disease evolution.

Foundational reference maps reshaped genomics by organizing genes, tissues, and cells into shared coordinate systems. We propose the phylophenome as the temporal counterpart for clinical medicine: a directed, biologically calibrated map of how human disease states succeed one another. The knowledge-base presented here is a first draft of it. Reading that knowledge-base as a set of clinical clades turns observational records into a coordinate system for studying disease evolution.

Transferring this approach to other EHR datasets from different regions across the world introduces new challenges. One challenge is the mapping to CCSR or ICD-10-CM. Even for datasets that are already encoded in different versions of the ICD-10 specific mapping tables to CCSR are required. For datasets enriched with SNOMED CT annotations, the official mapping tool [19] to the ICD-10-CM can be used to map to ICD-10-CM and then to CCSR [20]. Nevertheless, several of those datasets are only available in secured research environments adding additional challenges such as data ingress and egress, training, as well as limited available resources occur and require specialized solutions [21].

## 4 Methods

### 4.1 Data Source and Cohort Construction

De-identified EHR data were obtained from the Mass General Brigham (MGB) integrated healthcare system under institutional review board approval. Cohort construction followed the precision-phenotyping framework of Azhir et al.[16]. Briefly, three non-overlapping groups of adult patients (*≥*18 years) were assembled:

COVID-19 cases (n=85,364) with confirmed SARS-CoV-2 infection (positive test or recorded diagnosis), post-pandemic controls (n=170,497) with no recorded or probable SARS-CoV-2 infection, and pre-pandemic controls (n=39,817) drawn from the pre-2020 era. Controls were matched to cases on age, sex, race/ethnicity, region, and Charlson Comorbidity Index (CCI). Full inclusion and exclusion criteria, index-date definitions, and observation windows are detailed in Azhir et al. [16]. Baseline matching achieved balance on age (p>0.12), sex (p>0.45), and CCI (p=0.98, cases vs. post-pandemic controls).

### 4.2 Temporal Correlation Matrix Construction

Analogous to Azhir et al.[16], diagnoses were mapped to the Clinical Classifications Software Refined (CCSR)[14] taxonomy comprising 420 categories. For each ordered CCSR pair (*A, B*) within each patient, the dates of first occurrence of *A* and *B* were recorded and stratified into four temporal lag bins: 0–14 d (acute or concurrent), 15–30 d (subacute), 31–90 d (medium-term), and >90 d (chronic) using the tSPM+ algorithm [15]. Temporal precedence (*A* before *B*) was required.

For each phenotype pair and lag bin, the temporal association was quantified by Spearman’s rank correlation between the presence of *A* at the index time and the subsequent occurrence of *B* within the lag window, computed across all patients with complete observations:

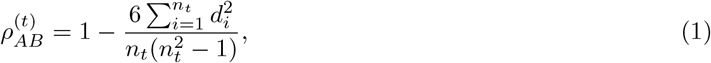

where *d*_*i*_ is the difference in ranks of patient *i* across the two binary outcomes within lag bin *t*, and *n*_*t*_ is the number of patients with informative observations in that bin. Statistical significance was assessed by permutation test (10,000 permutations of the target label vector with the source held fixed), with *p*-values adjusted by the Benjamini-Hochberg false discovery rate procedure at *α*=0.05 [22]. This yielded a 420 × 420 × 4-bin reference corpus, reduced to 435,240 rows after pruning of empty cells and self-loops.

### 4.3 Annotation Provenance and Label Distillation

Because manual chart review cannot scale to half a million sequences, we automated the initial adjudication through a two-stage protocol. In the first stage, a Labeling Function Development Set of 200 phenotype pairs was assessed using online biomedical literature. Two reviewers independently recorded a preliminary binary label (noise or non-noise) and a brief mechanistic rationale, with disagreements resolved by discussion. In the second stage, we deployed the MedGemma-7B model on local CLAI group servers to enforce strict data privacy. Inference was executed between March and April 2026. Decoding parameters were set to a temperature of 0.0 to enforce deterministic outputs, with a maximum generation limit of 4800 tokens. Iterative manual prompt engineering used the 200 human-labeled examples to refine prompt structure, instruction clarity, and decision criteria. Inter-method agreement between the final MedGemma labels and the human-consensus reference on the 200-row development set was quantified by Cohen’s *κ*. Once calibrated, the prompt generated silver-standard labels for a training corpus of 10,000 distinct clinical sequences.

### 4.4 Feature Extraction

Features were extracted from the full 435,240-row reference corpus and joined to the labeled subset by [`sequence, duration`] to prevent data leakage. The two classifiers received partially distinct feature sets, reflecting a deliberate architecture-driven design rather than an arbitrary asymmetry. A shared five-class foundation was computed for both models. Class 1 captured core statistical signal: Spearman *ρ, ρ*^2^, −log_10_(*p*_adj_), signal strength (|*ρ*|× −log_10_(*p*_adj_)), and a log Bayes factor. Class 2 captured directional asymmetry: reverse-direction |*ρ*|, directional dominance, and direction confidence. Class 3 captured cross-duration temporal profile: temporal slope across lag bins, within-pair *z*-score relative to the pair’s own cross-lag distribution, temporal AUC, coefficient of variation, temporal range, and lag-interaction terms. Class 4 captured corpus-relative empirical priors: source and target fan-out, *z*-scores versus source- and target-level *ρ* distributions, and association surprise. Class 5 captured clinical ontology: primary body-system identity from the CCSR-to-systems mapping, data-driven system-to-system relatedness, phenotype specificity score, and acute/chronic keyword flags. All graph features were computed on a directed reference graph built from the full corpus at *p*_adj_ < 0.05, with edge weights *w* = −log_10_(*p*_adj_) × |*ρ*| .

The models then diverged in their higher-order features, by design. The Random Forest has no native mechanism for multi-hop structural reasoning, so two additional feature classes were computed to supply this information as tabular summaries: Classes 7–8 added one- and two-step Markov transition probabilities, normalised pointwise mutual information (NPMI), HITS hub and authority scores, shared downstream Jaccard coefficient, target PageRank, and target betweenness centrality. The GNN did not receive these classes because its two-layer TransformerConv message-passing encodes equivalent structural information directly from the graph; supplying the same quantities as additional edge features would introduce redundancy without informational gain. Conversely, the GNN received richer versions of the shared classes, exploiting its attention mechanism’s ability to weight fine-grained local signal per neighbourhood context: Class 1 was extended with a Bayesian posterior probability term and a *p*_adj_ < 0.001 indicator; Class 2 with sign concordance, both-directions significance, and asymmetric bidirectionality flags; Class 3 with observed-window counts, proportion of significant duration bins, and early-peak, late-peak, and plateau pattern flags; Class 4 with per-duration *z*-scores and source selectivity; and Class 5 with procedure-to-diagnosis transition type flags and a systemic-to-specific indicator. Class 6, unique to the GNN, captured network position features used to construct node representations for message passing: target PageRank, betweenness centrality, transition entropy, local transition probability, cascade potential, reciprocal edge indicator, and an `edge_in_reference` flag that defined the message-passing subgraph. Because the RF and GNN are deployed as complementary components of a late-fusion average (*P*_avg_ = (*P*_RF_ + *P*_GNN_)*/*2) rather than as direct competitors, the asymmetric feature design optimises each model for its architectural role; the averaged output, not the individual classifiers, is the unit evaluated against independent genetic evidence.

### 4.5 Random Forest Classifier

The 10,000-row labeled training corpus was split into training (80%) and held-out test (20%) partitions using stratified sampling. Near-zero-variance features were identified on the training partition alone and removed from both partitions prior to model fitting. The RF was trained using the MLHO framework [23]. The Youden-optimal decision threshold was selected from the training ROC curve 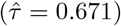. The trained model was subsequently applied to the 200-row development set using the human-adjudicated label as the reference standard.

### 4.6 Graph Neural Network Classifier

The GNN implemented a two-layer TransformerConv architecture [24] for directed edge classification. Node representations were constructed by aggregating source-node and target-node features (fan-out, PageRank, betweenness centrality, transition entropy, phenotype specificity, and acute/multisystem flags) from the full corpus via grouped means. Message passing used a minimal five-feature edge representation (*ρ*, duration ordinal, signal strength, forward-only indicator, posterior probability) to prevent feature shortcutting during neighbourhood aggregation. Edge classification concatenated source node embedding, destination node embedding, and the full 37-feature edge representation, passing the result through a three-layer MLP with batch normalisation and dropout at each layer. Hyperparameters were selected by exhaustive grid search over hidden dimension ∈ {64, 128}, dropout ∈ {0.3, 0.5}, weight decay ∈ {10^−3^, 10^−4^}, label smoothing ∈ {0.0, 0.05}, and edge-feature dropout ∈ {0.0, 0.1}, with learning rate fixed at 10^−3^ and two attention heads across all combinations. The winning configuration was hidden dim = 128, dropout = 0.3, weight decay = 10^−4^, label smoothing = 0.05, edge-feature dropout = 0.0. The final model was trained with AdamW [25], cosine annealing learning rate schedule (minimum 10^−5^), and gradient clipping (maximum norm = 1.0), for up to 200 epochs with early stopping after 40 epochs without improvement on test AUROC. Training used a class-weighted binary cross-entropy loss with automatic positive weight = *n*_neg_/*n*_pos_ and label smoothing = 0.05. Reproducibility was ensured by `set_seed`(42) applied before both grid search evaluation and final training. The message-passing graph comprised all corpus associations meeting *p*_adj_ < 0.05 (117,697 directed edges).

### 4.7 UKBB–CCSR Phenotype Mapping and Genetic Correlation Attachment

Genome-wide genetic correlation estimates (*r*_*g*_) were obtained from the Neale Lab LD score regression (LDSC) browser (https://ukbb-rg.hail.is), which provides pairwise genetic correlation estimates derived from genome-wide association study summary statistics in approximately 361,000 unrelated white British participants from the UK Biobank [12, 26]. To align UK Biobank disease traits with the Clinical Classifications Software Refined (CCSR) phenotype taxonomy used in the TKB, a two-tier ICD-10 mapping strategy was implemented. UK Biobank traits with explicit ICD-10 diagnoses embedded in their labels were mapped directly, whereas remaining disease traits were manually harmonised to three-character ICD-10 categories through systematic review of UK Biobank phenotype documentation. Non-clinical traits (e.g., biomarkers, imaging, and dietary variables) were excluded.

CCSR phenotype categories were linked to UK Biobank traits through their constituent ICD-10-CM billing codes. Because LDSC-derived genetic correlations are indexed at the three-character ICD-10 resolution, ICD-10-CM codes were truncated to their corresponding three-character ICD-10 category (i.e., the World Health Organization base disease code). A phenotype pair was considered mappable when both source and destination phenotypes resolved to at least one UK Biobank disease trait. Where multiple UK Biobank trait combinations were available for a single phenotype pair, the *r*_*g*_ estimate with the lowest *p*-value was retained to prioritise the strongest statistically supported signal. Phenotype pairs without an available genetic correlation estimate were excluded from genomic validation analyses.

### 4.8 Genetic Validation

Classifier confidence was evaluated against independent genomic evidence from the UK Biobank by assessing whether high-confidence temporal trajectories preferentially aligned with shared genetic architecture. Phenotype pairs were classified as genetically supported when they exhibited both a statistically significant genetic correlation 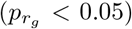 and an effect of at least moderate magnitude (|*r*_*g*_ |> 0.15). The dual criterion required supported pairs to show not merely a detectable signal but a substantive degree of shared heritability, reducing the contribution of weak correlations that reach significance largely by virtue of the UK Biobank’s large sample size. To avoid inflation arising from multiple CCSR phenotypes mapping to the same UK Biobank trait combination, one representative row per unique *r*_*g*_ estimate was retained.

Classifier probabilities were stratified into ten confidence deciles and the prevalence of genetically supported phenotype pairs was computed within each stratum. Top-decile enrichment relative to the corpus-wide baseline was assessed by permutation testing (1,000 iterations; shuffled-label null). Because the mappable subset is restricted to heritable common diseases and resolved at coarser diagnostic granularity than EHR data, *r*_*g*_ was treated as an orthogonal plausibility signal rather than ground truth, and decile enrichment as a within-subset ranking rather than a population prevalence estimate.

### 4.9 Unsupervised Mechanistic Discovery

Two complementary discovery protocols were applied to the full adjudicated sequence matrix. For each protocol, an unweighted late-fusion average *P*_avg_ = (*P*_RF_ + *P*_GNN_)/2 was computed from the full all-corrs prediction outputs; tSPM *ρ* and *p*-values were joined from the upstream correlation file by [`sequence, duration`]; UKBB 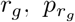, and *r*_*g*,SE_ were joined from the full genetic correlation file by [`sequence, duration`]. No stacking weights were learned for the late-fusion average and the unweighted average is the variance-minimising combination under uncertainty about the relative calibration of the two models.

**Mechanical cascade identification** isolated progressions operating independently of shared polygenic susceptibility. Sequences were filtered for extreme joint confidence (*P*_avg_ > 0.95) combined with strictly confirmed null genomic correlation 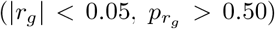. Rows without a UKBB genetic correlation entry were excluded; a measured null was required, not a missing value. This combination structurally isolates acquired, iatrogenic, or post-acute progressions from pleiotropic effects.

**Topological bridge identification** exploited systematic divergence between the two constituent models. Sequences were filtered for low RF probability (*P*_RF_ < 0.40) combined with high GNN probability (*P*_GNN_ > 0.90). Because the RF operates on tabular local edge features while the GNN aggregates neighbourhood topology via message passing, divergence in this direction identifies latent clinical pathways that are sparse in direct temporal co-occurrence but are structurally enforced by the surrounding phenotype network. No genetic filter was applied; model divergence itself is the discovery criterion.

## 5 Conclusion

We have presented a biologically calibrated, directed map of disease progression, built from longitudinal electronic health records and validated against orthogonal genetics. It reads observational records as a temporal coordinate system rather than a static network, and it stands as a first instantiation of the phylophenome. The current version carries one clear boundary. It is built entirely from New England, so its trajectories reflect one region’s population, its care patterns, and its coding conventions. We treat that boundary as both a limitation and a direction for the work that follows.

We will compute the same corpus from other regions and compare their progression maps directly. The marginal cost of curating an additional regional corpus is small against the value of that comparison, because the method and the pipeline already exist. Planned sources include the UK Biobank, the All of Us research program, Statistics Denmark, and insurance coverage data, which together extend the map toward national and cross-national scale. Where regions diverge, we read the divergence as signal about environment, access, and clinical practice, not as noise to average away. A reference map that resolves those differences will say more about human disease than any single population can reveal on its own.

## Data Availability

Raw EHR data are restricted by MGB institutional policy to protect patient privacy. The adjudicated knowledge-base edge-list and classifier probability scores are provided in Supplementary Data 1.

## Ethics and Declarations

This study was approved by the Mass General Brigham Institutional Review Board. Patient consent was waived due to the use of de-identified administrative data for retrospective analysis.

## Data and Code Availability

Raw EHR data are restricted by MGB institutional policy to protect patient privacy. The adjudicated knowledge-base edge-list and classifier probability scores are provided in Supplementary Data 1. The analytical pipeline, extraction algorithms, model weights, MedGemma prompt, and figure-generation scripts are publicly available via the CLAI research group GitHub repository (https://github.com/clai-group/claid-infrastructure). An interactive browser for the knowledge-base, ClaiD, is available at https://clai-group.github.io/claid-infrastructure/. Readers can filter trajectories by confidence, time window, and genetic support, then assemble and export clinical clades.

